# Antimicrobial Susceptibility Patterns in Adult Patients Hospitalized for Community-acquired Urinary Tract Infection in Tertiary Hospitals in Chile

**DOI:** 10.1101/2022.12.27.22283981

**Authors:** Luis Rojas, Inés Ceron, Esteban Araos-Baeriswyl, Paola Viviani, Rodrigo Olivares, Andrés Valenzuela, Andrés Aizman

## Abstract

**Background:** The constant increase of factors associated with the presence of resistant strains makes empirical antibiotic selection a challenge in patients hospitalized for community-acquired urinary tract infection. We characterized the type of bacteria and their antimicrobial susceptibility pattern in cultures obtained from adult patients that needed hospitalization for this disease in two tertiary hospitals in Chile.

**Methods:** We conducted a cross-sectional study in adults hospitalized for community-acquired urinary tract infection between 2017 and 2022. A total of 830 patients were included. All patients with positive cultures were included in the analysis.

**Results:** Escherichia coli was the most frequent infectious agent (68.1%), followed by Klebsiella spp. (17.7%) and Enterococcus faecalis (6.7%). Among Enterobacteriaceae strains, 35.2%, 19.7% and 27.2% were resistant to first, second and third-generation cephalosporin, respectively. 36.9% were resistant to ciprofloxacin and 1.8% to amikacin. Gram-positive bacteria were resistant to oxacillin and ampicillin in 25% and 18%, respectively.

**Conclusion:** We detected a high prevalence of community-acquired urinary tract infections caused by bacteria resistant to commonly used antibiotics in adult patients that need to be hospitalized. In view of this, we believe that current medical guidelines should be reviewed and updated.

## INTRODUCTION

Community-acquired urinary tract infection (CA-UTI) is one of the most frequent bacterial infections in the community and hospitalized patients (1,2). Most are caused by gram-negative enterobacteria, with Escherichia coli being the bacterium detected in more than 50% of cases (3). Their initial treatment is based on the empirical use of antibiotics for at least 48 hrs, until antimicrobial susceptibility studies are obtained (4). This initial empirical antibiotic selection is very important, because if it is inadequate, increases morbidity, costs, and length of stay in the hospital (5,6).

Local reports of antimicrobial susceptibility patterns in patients with community or hospital-acquired urinary tract infections provide important support for the selection of antibiotic treatment before receiving culture results (4). However, this information should be constantly updated due to the significant and constant increase in risk factors associated with the presence of resistant strains (7–11). The information should also specify whether samples correspond to ambulatory patients or those who required hospitalization for CA-UTI, the latter being a group at greater risk of suffering consequences due to an inadequate initial selection of antibiotics (12–15).

In view of the above, in addition to the scarce information published on the subject in Chile and Latin America, the aim of this study is to characterize the type of bacteria and their antimicrobial susceptibility pattern in cultures obtained from patients hospitalized for CA-UTI, with a view to determining the prevalence of bacteria resistant to first-line antibiotics.

## MATERIALS AND METHODS

This was a cross-sectional study that incorporated prospective sampling in two hospitals/health centers within the Metropolitan Region of Chile: UC Hospital Clinic (HCUC) and Dr. Sotero del Rio Hospital (HSR), belonging to private and public health system, respectively. All patients over 18 years of age hospitalized for CA-UTI were recruited, in consecutive order. Patients included those who presented a UTI at the time of hospital admission, and those who developed UTI within the first 48 hours of hospitalization.

Patients not included in the sample were those with recent hospital stays (7 days prior), polymicrobial cultures, cultures from urinary catheter users without previous catheter change, fungal cultures, cultures from urine collection bags, cultures from patients with ileal reservoir, and positive cultures without antimicrobial susceptibility study.

The diagnosis of CA-UTI was based on the presence of suggestive symptoms (dysuria, pollakiuria, urinary incontinence, lumbar or flank pain, fever, chills and/or acute changes in mental status) associated to urine sediment with leukocyturia (> 10 leukocytes per field). Growth of a uropathogen in a urine culture was greater than or equal to 105 colony forming units (cfu)/mL from spontaneous urination samples, or greater than or equal to 103 cfu/mL if collected through bladder catheterization, urinary catheter puncture and/or bladder puncture (16,17). Urine samples were obtained by spontaneous urination of second stream urine, bladder catheterization, urinary catheter puncture and/or bladder puncture. Seeding was performed on chromogenic agar (CHROMID© CPS© Elite / Columbia CNA + 5% sheep blood Agar, bioMerieux) with 1 μL calibrated loop for second stream urine, 10 μL calibrated loop for catheterizations and urinary catheter puncture, and 100 μL for bladder puncture. Incubation was performed at 35°C for 16 hours in an aerobic environment. After incubation, colony counting and bacterial identification was performed by mass spectrometry (Matrix-Assisted Laser Desorption/Ionization Time-Of-Flight MALDI-TOF) in the Microflex© spectrometer, using the Biotyper© system (Bruker) and following the manufacturer’s instructions. Antimicrobial susceptibility of Escherichia coli was performed on the automated VITEK 2 XL system© (bioMerieux) with the N-250 card (bioMerieux). The rest of the bacteria was analyzed by agar dilution, using the Cathra© multiplier (MCT Medical Inc. St. Paul, Minn.). The antibiotics included are as follows: nitrofurantoin, amikacin, gentamicin, cephalosporins, trimethoprim-sulfamethoxazole, ciprofloxacin, oxacillin and ampicillin (first-line antibiotics), piperacillin-tazobactam, carbapenems, cefoperazone-sulbactam, vancomycin, teicoplanin and linezolid (second-line antibiotics). The interpretation of the minimum inhibitory concentration (MIC) was performed for each antimicrobial and bacteria type, according to *Clinical and Laboratory Standards Institute* (CLSI) (18).

Data were obtained from clinical and laboratory service records, and were entered anonymously in a database implemented through the RedCap system.

## STATISTICS

Categorical variables were expressed as frequencies and percentages, and numeric variables as medians and range, with a 95% confidence interval (CI 95%). The association between categorical variables was established using Fisher’s exact test or Chi-square test as appropriate. An *a priori* alpha value of 5% was defined to establish statistically significant differences. The analyses were performed using the STATA v. 14 software.

This study was approved by the Health Sciences Scientific Ethics Committee of the School of Medicine of the Pontifical Catholic University and the South-East Metropolitan Health Service (ID 2003005003).

## RESULTS

A total of 830 patients were recruited between October 2017 and September 2021, of which 88.7% were recruited at the university’s hospital clinic (HCUC) (n = 736). The mean age was 66.5 (range: 18 to 102 years), 64.1% were female. All CA-UTI were diagnosed with urine cultures, 16.3% of all cases had concomitant positive blood cultures.

The most frequent comorbidities within the sample were diabetes mellitus (33.9%) and chronic renal insufficiency (21.4%). Of all patients, 43.3% presented at least one previous hospitalization during the last year. An indwelling urinary catheter had been used by 13.3% of patients at least 90 days prior to admission (mean days of use = 51.3, range = 1 to 365 days), of which 35.5% were using it at the time of admission. Some 28.3% presented anatomical or functional alterations of the urinary tract, and 14.1% had undergone instrumentalization of the urinary tract during the last year. About 24.5% had previous cultures for resistant bacteria in the last year, and 45.7% had used antibiotics in the last 90 days prior to admission. Regarding the severity of the clinical scenario, most of the patients (60.5%) were admitted to low complexity hospital units. The average length of stay was 10.5 days, the median was 7 days, and 5.1% died during their stay **(Table 1)**.

**Table 1.**
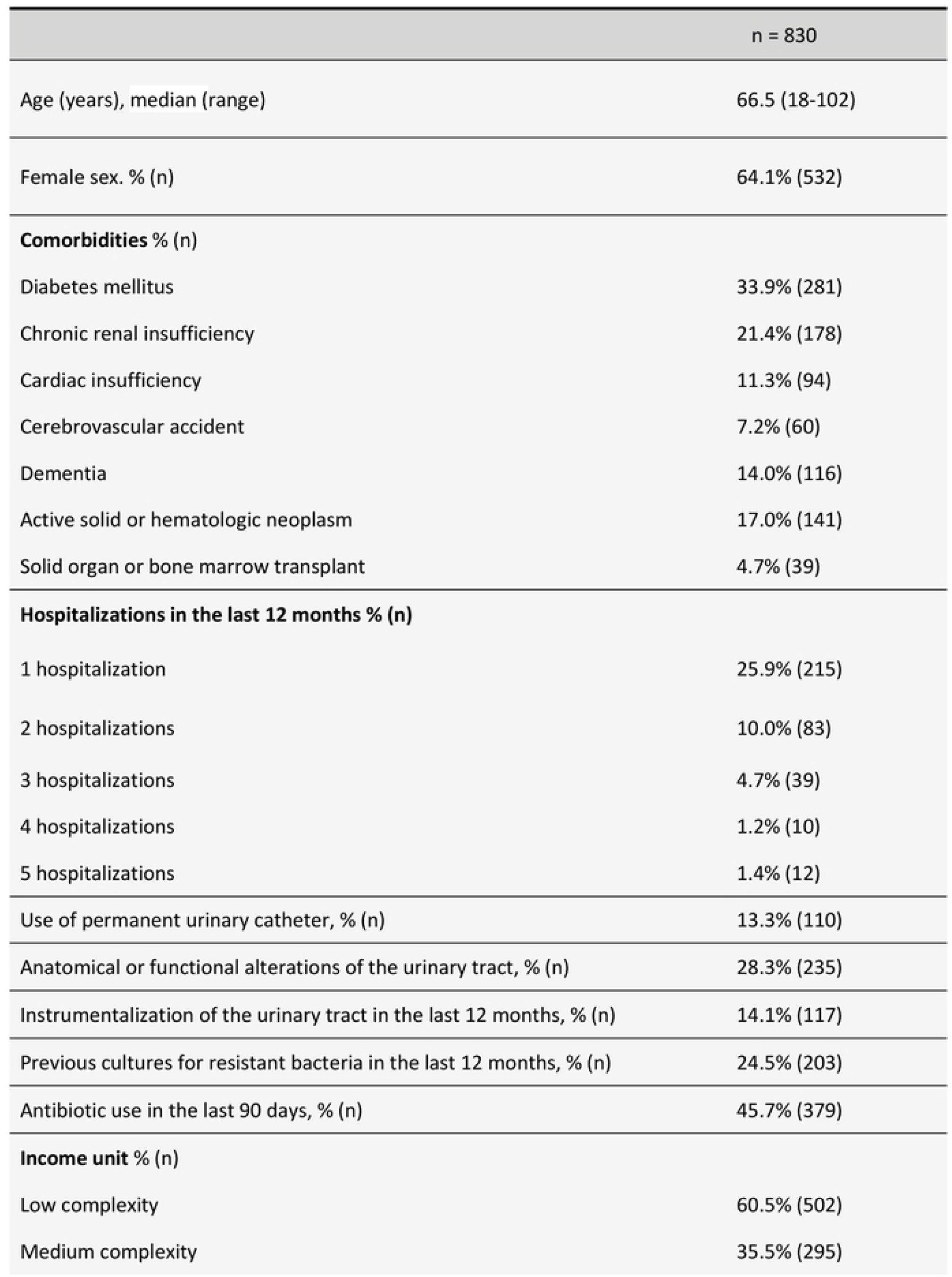
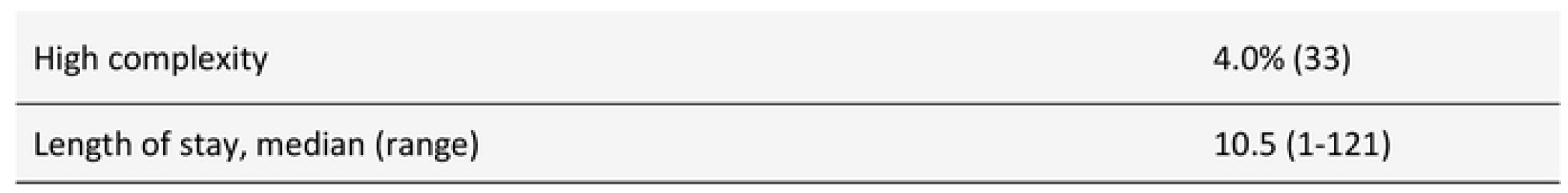
Clinical and demographic characteristics of the patients participating in the study.

### Cultures

Enterobacteriaceae were the most frequent infectious agents identified. *Escherichia coli* was found in 68.1% of cultures, *Klebsiella spp*. in 17.7%, *Proteus spp*. 4.6% and *Pseudomonas aeruginosa* 2.5%. Gram-positive pathogens were detected in 88 patients, including *Enterococcus faecalis* (6.7%), *Enterococcus faecium* (1.9%) and *Staphylococcus spp*. (1.9%). The distribution was similar between both hospitals/healthcare centers included in the study, except for *Klebsiella spp*, which was more frequent in the Dr. Sotero del Rio Hospital (HSR) **(table 2)**.

**Table 2.** Type of bacteria isolated in urine cultures, showing differences according to care center.

|  | Both centers (n=830) | HCUC (n=736) | HSR (n=94) | p |
| --- | --- | --- | --- | --- |
| Bacterial isolate | % (n) | % (n) | % (n) |  |
| *Escherichia coli* | 68.1% (565) | 69.0% (508) | 60.6% (57) | p<br>>0.05 |
| *Klebsiella spp.* | 17.7% (147) | 16.6% (122) | 26.6% (25) | **p**<br>**0.02** |
| *Enterococcus faecalis* | 6.7% (56) | 7.2% (53) | 3.2% (3) | p<br>>0.05 |
| *Proteus spp.* | 4.6% (38) | 5.0% (37) | 1.1% (1) | p<br>>0.05 |
| *Pseudomonas aeruginosa* | 2.5% (21) | 2.6% (19) | 2.1% (2) | p<br>>0.05 |
| *Enterococcus faecium* | 1.9% (16) | 1.8% (13) | 3.2% (3) | p<br>>0.05 |
| *Staphylococcus spp.* | 1.9% (16) | 1.8% (13) | 3.2% (3) | p<br>>0.05 |
| *Enterobacter cloacae (complex)* | 1.1% (9) | 1.1% (8) | 1.1% (1) | p<br>>0.05 |
| Other Enterobacteriaceae* | 1.1% (9) | 1.2% (9) | 0.0% (0) | p<br>>0.05 |
| Other bacteria** | 0.7% (6) | 0.8% (6) | 0.0% (0) | p<br>>0.05 |

\* The following are grouped under "Other Enterobacteriaceae": *Morganella morganii* (n: 4), *Citrobacter spp.* (n: 3), *Serratia marcescens* (n: 2).
\*\* The following are grouped under "Other Bacteria": *Streptococcus agalactiae group B* (n: 2), *Streptococcus gallolyticus/equinus complex* (n: 1), *Enterococcus gallinarum* (n: 1), *Enterococcus avium* (n: 1) and *Achromobacter spp.* (n: 1).

A lower prevalence of *Escherichia coli* was observed in male population compared to female population (51.7% vs 77.3%), with the opposite occurring for *Klebsiella spp* (24.2% vs 14.1%), *Enterococcus faecalis* (10.1% vs 4.9%), *Proteus spp* (7% vs 3.2%) and *Pseudomonas aeruginosa* (4.7% vs 1.3%). In turn, those older than 65 had a higher incidence of *Enterococcus faecalis* (8.2% vs 4.5%) and *Pseudomonas aeruginosa* (3.6% vs 0.9%) (**Table 3)**.

**Table 3.**
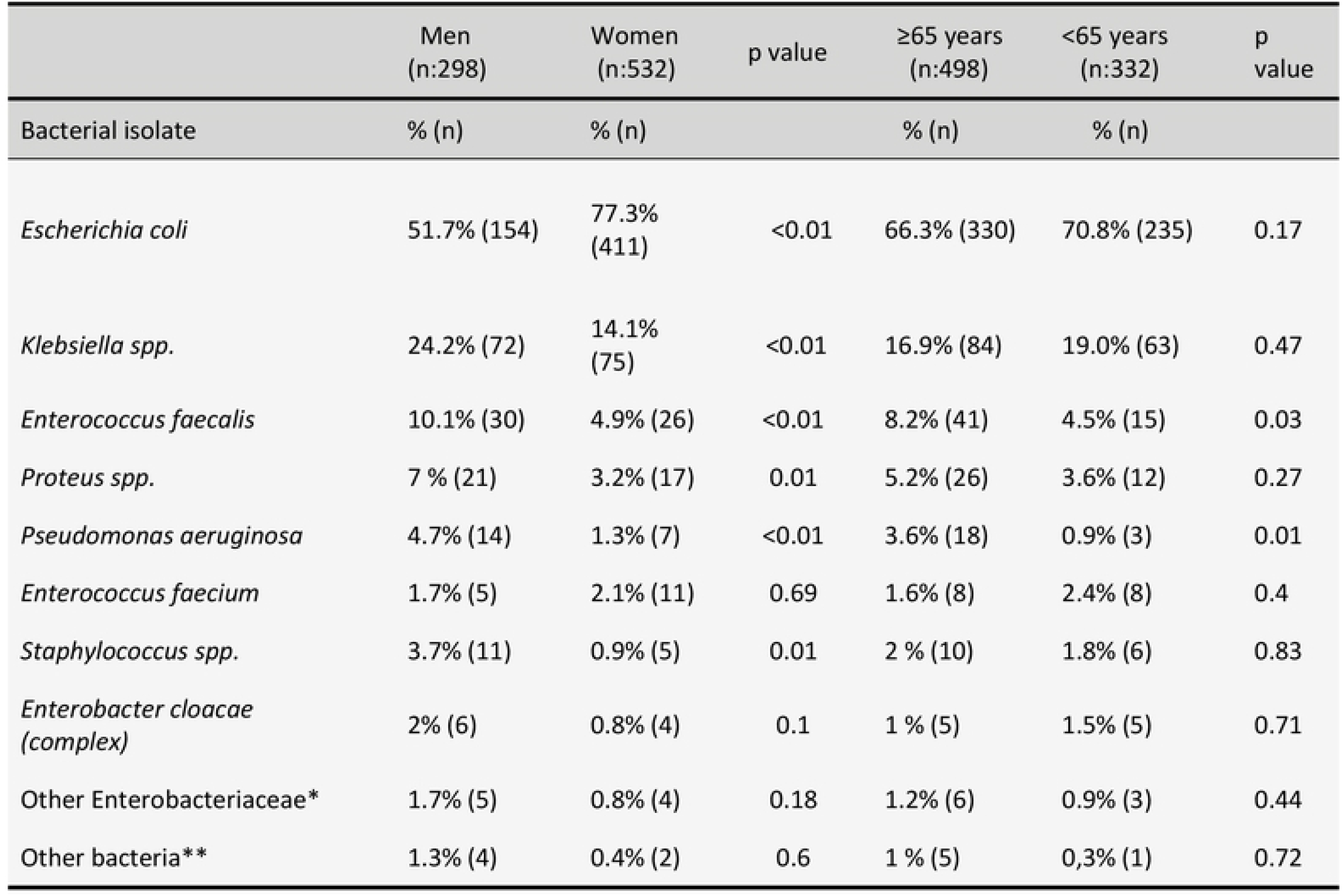
Type of bacteria isolated in urine cultures, showing differences by sex and age group in both recruiting centers

|  | Men<br>(n:298) | Women<br>(n:532) | p value | ≥65 years<br>(n:498) | <65 years<br>(n:332) | p<br>value |
| --- | --- | --- | --- | --- | --- | --- |
| Bacterial isolate | % (n) | % (n) |  | % (n) | % (n) |  |
| *Escherichia coli* | 51.7% (154) | 77.3%<br>(411) | <0.01 | 66.3% (330) | 70.8% (235) | 0.17 |
| *Klebsiella spp.* | 24.2% (72) | 14.1%<br>(75) | <0.01 | 16.9% (84) | 19.0% (63) | 0.47 |
| *Enterococcus faecalis* | 10.1% (30) | 4.9% (26) | <0.01 | 8.2% (41) | 4.5% (15) | 0.03 |
| *Proteus spp.* | 7 % (21) | 3.2% (17) | 0.01 | 5.2% (26) | 3.6% (12) | 0.27 |
| *Pseudomonas aeruginosa* | 4.7% (14) | 1.3% (7) | <0.01 | 3.6% (18) | 0.9% (3) | 0.01 |
| *Enterococcus faecium* | 1.7% (5) | 2.1% (11) | 0.69 | 1.6% (8) | 2.4% (8) | 0.4 |
| *Staphylococcus spp.* | 3.7% (11) | 0.9% (5) | 0.01 | 2 % (10) | 1.8% (6) | 0.83 |
| *Enterobacter cloacae*<br>(complex) | 2% (6) | 0.8% (4) | 0.1 | 1 % (5) | 1.5% (5) | 0.71 |
| Other Enterobacteriaceae* | 1.7% (5) | 0.8% (4) | 0.18 | 1.2% (6) | 0.9% (3) | 0.44 |
| Other bacteria** | 1.3% (4) | 0.4% (2) | 0.6 | 1 % (5) | 0,3% (1) | 0.72 |

### Antimicrobial Resistance

#### Escherichia coli

Resistance rates for 1st generation cephalosporins, trimethoprim-sulfamethoxazole and ciprofloxacin exceeded 30%, and for 2nd and 3rd generation cephalosporins resistance rates were 15% and 20.5%, respectively. Resistance to nitrofurantoin was observed in only 3.9% of cases, being even lower for piperacillin-tazobactam and carbapenems (less than 1%). The same was observed for amikacin, but not for gentamicin (0.2% and 11.3%, respectively). **(Table 4)**.

**Table 4.** Antibiotic resistance pattern for total bacteria found in urine cultures of patients with community-acquired urinary tract infection admitted to HCUC or HSR. N/A: not applicable

| Antibiotic | *Escherichia coli*<br>(n: 565); % (n) | *Klebsiella spp.*<br>(n:147); % (n) | *Proteus spp.*<br>(n:38); % (n) | *Enterobacter cloacae*<br>(n:9); % (n) | *Pseudomona aeruginosa*<br>(n:21); % (n) | Enterococcus faecalis<br>(n:56); % (n) | Enterococcus faecium<br>(n:16); % (n) | Staphylococcus spp.<br>(n:16); % (n) | Total; % (n) |
| --- | --- | --- | --- | --- | --- | --- | --- | --- | --- |
| Cephalosporins 1<sup>st</sup> | 31.9% (180) | 53.1% (78) | 15.8% (6) | N/A | N/A | N/A | N/A | N/A | 35.2% (264) |
| Cephalosporins 2<sup>nd</sup> | 15.0% (85) | 39.5% (58) | 13.1% (5) | N/A | N/A | N/A | N/A | N/A | 19.7% (148) |
| Nitrofurantoin | 3.9% (22) | 32.7% (48) | 36.8% (33) | 44.4% (4) | N/A | 0.0% (0) | 68.8% (11) | 0.0% (0) | 13.9% (118) |
| Ciprofloxacin | 35.2% (199) | 44.2% (65) | 18.4% (7) | 33.3% (3) | 28.6% (6) | 37.5% (21) | 81.3% (13) | N/A | 36.9% (314) |
| Gentamicin | 11.3% (64) | 32.7% (48) | 5.3% (2) | 22.2% (2) | 19.0% (4) | N/A | N/A | 12.5% (2) | 15.3% (122) |
| Amikacin | 0.2% (1) | 6.8% (10) | 2.6% (1) | 22.2% (2) | 0.0% (0) | N/A | N/A | N/A | 1.8% (14) |
| Trimethoprim-Sulfamethoxazole | 32.7% (185) | 44.2% (65) | 21.1% (8) | 44.4% (4) | N/A | N/A | N/A | 12.5% (2) | 34.1% (264) |
| Cephalosporins 3<sup>rd</sup> | 20.5% (116) | 53.1% (78) | 18.4% (7) | 44.4% (4) | 33.3% (7) | N/A | N/A | N/A | 27.2% (212) |
| Cefoperazone-Sulbactam | N/A | N/A | N/A | N/A | 0.0% (0) | N/A | N/A | N/A | 0.0% (0) |
| Piperacillin-tazobactam | 0.7% (4) | 17.7% (26) | 2.6% (1) | 44.4% (4) | 4.8% (1) | N/A | N/A | N/A | 4.6% (36) |
| Ertapenem | 0.6% (3) | 17.7% (26) | 0.0% (0) | 33.3% (3) | N/A | N/A | N/A | N/A | 4.2% (32) |
| Imipenem | 0.2% (1) | 3.4% (5) | 0.0% (0) | 11.1% (1) | 33.3% (7) | N/A | N/A | N/A | 1.8% (14) |
| Meropenem | 0.0% (0) | 4.1% (6) | 0.0% (0) | 0.0% (0) | 14.3% (3) | N/A | N/A | N/A | 1.2% (9) |
| Ampicillin | N/A | N/A | N/A | N/A | N/A | 0.0% (0) | 81.3% (13) | N/A | 18% (13) |
| Oxacillin | N/A | N/A | N/A | N/A | N/A | N/A | N/A | 25.0% (4) | 25.0% (4) |
| Vancomycin | N/A | N/A | N/A | N/A | N/A | 0.0% (0) | 56.3% (9) | 0.0% (0) | 10.2% (9) |
| Teicoplanin | N/A | N/A | N/A | N/A | N/A | 0.0% (0) | 6.3% (1) | N/A | 1.4% (1) |
| Linezolid | N/A | N/A | N/A | N/A | N/A | 0.0% (0) | 0.0% (0) | N/A | 0.0% (0) |

#### *Klebsiella* spp

Except for amikacin (6.8%), all 1st line antibiotics presented resistances above 30%. The highest rates were observed for 1st and 3rd generation cephalosporins, both with a 53.1% resistance, then for ciprofloxacin and trimethoprim-sulfamethoxazole (both 44.2%), and lastly for nitrofurantoin and gentamicin (both 32.7%).

With respect to 2nd line antibiotics, less resistance was detected, 17.7% was found for ertapenem and piperacillin-tazobactam, 3.4% for imipenem, and 4.1% for meropenem. Of the latter (11 strains), three included carbapenemases (KPC), and two consisted of New Delhi metallo-beta-lactamase **(NDM)** carbapenemases. **(Table 4)**.

#### *Proteus* spp

*Proteus spp*. presented resistance rates between 13% and 21% for 1st and 2nd generation cephalosporins, ciprofloxacin and trimethoprim-sulfamethoxazole. Similar resistance values were observed for 3rd generation cephalosporins (18.4%). In contrast to previous cases, a high resistance to nitrofurantoin was identified (86.8%), the highest observed in all the cultures recorded. Regarding aminoglycosides, 6.8% and 32.7% resistance was observed for amikacin and gentamicin, respectively. No strains resistant to carbapenems were detected, and only 2.6% were resistant to piperacillin-tazobactam **(Table 4)**.

#### Enterobacter Cloacae (complex)

Of the 9 strains detected, 33% were resistant to ciprofloxacin, 44% to trimethoprim-sulfamethoxazole, nitrofurantoin and 3rd generation cephalosporins. They had higher resistance rates for aminoglycosides (22.2% for amikacin and gentamicin), piperacillin-tazobactam (33.3%) and ertapenem (33.3%). One strain showed resistance to imipenem, which was through the production of VIM carbapenemases **(Table 4)**.

#### Pseudomona aeruginosa

Of all the strains, 28.6% and 33.3% were resistant to ciprofloxacin and ceftazidime, respectively. Together with ***Escherichia coli*** strains, they presented the lowest resistance rates for piperacillin-tazobactam and amikacin (4.8% and 0%, respectively). In turn, the resistance observed for carbapenems was the highest (33.3% for imipenem and 14.3% for meropenem). None of them were carbapenemases producers **(Table 4)**.

#### Enterococcus faecalis and Enterococcus faecium

Within the *Enterococcus faecalis* strains, 37.5% were resistant to ciprofloxacin and all were sensitive to ampicillin. Of the *Enterococcus faecium* strains, 81.3% were resistant to ampicillin and ciprofloxacin, and 56.3% to vancomycin. They were all sensitive to linezolid **(Table 4)**.

#### Staphylococcus spp

Within this group, composed of ***Staphylococcus aureus*** (9), ***Staphylococcus*** epidermidis (3), ***Staphylococcus*** haemolyticus (2) and ***Staphylococcus*** lugdunensis (2), 25% were resistant to oxacillin and all cases were sensitive to vancomycin **(Table 4)**.

### Comparison of urine cultures according to origin and demographic variables

When comparing the prevalence of microorganisms from both healthcare centers, there were only differences for *Klebsiella spp*. (HCUC 16.6% and HSR 26.6%) (Table 2). Regarding antimicrobial susceptibility patterns for the two most frequent types of bacteria (i.e., ***Escherichia coli and*** *Klebsiella spp*), greater resistance was observed for almost all antibiotics in the strains from the HSR, especially for 1st generation cephalosporins, piperacillin-tazobactam, nitrofurantoin and ertapenem. **(Table 5)**.

**Table 5.** Antimicrobial resistance profile for *E. coli* and *Klebsiella spp*. in each hospital center, antibiograms reported from urine cultures.

| Antibiotic | *Escherichia Coli* |  |  | *Klebsiella spp* |  |  |
| --- | --- | --- | --- | --- | --- | --- |
|  | HCUC<br>(n:508); % (n) | HSR<br>(n:57); %<br>(n) | *p* | HCUC<br>(n:122); %<br>(n) | HSR<br>(n:25); %<br>(n) | *p* |
| Cephalosporins 1<sup>st</sup> | 30.3% (154) | 45.6% (26) | 0.018 | 50.8% (62) | 64.0% (16) | 0.01 |
| Cephalosporins 2<sup>nd</sup> | 15.0% (85) | N/A | - | 47.5% (58) | N/A | - |
| Nitrofurantoin | 3.3% (17) | 8.8% (5) | 0.04 | 29.5% (36) | 48.0% (12) | 0.22 |
| Ciprofloxacin | 34.3% (174) | 43.9% (25) | 0.14 | 41.0% (50) | 60.0% (15) | 0.07 |
| Gentamicin | 12.0% (61) | 5.3% (3) | 0.12 | 31.1% (38) | 40.0% (10) | 0.08 |
| Amikacin | 0.2% (1) | 0.0% (0) | 0.18 | 6.6% (8) | 8.0% (2) | 0.86 |
| Trimethoprim-sulfamethoxazole | 31.9% (162) | 40.4% (23) | 0.19 | 43.4% (53) | 48.0% (12) | 0.38 |
| Cephalosporins 3<sup>rd</sup> | 20.3% (103) | 22.8% (13) | 0.65 | 53.3% (65) | 52.0% (13) | 0.9 |
| Piperacillin-Tazobactam | 0.8% (4) | 7.0% (4) | <0.01 | 12.3% (15) | 44.0% (11) | <0.01 |
| Ertapenem | 0.4% (2) | 1.8% (1) | 0.7 | 13.9% (17) | 36.0% (9) | <0.01 |
| Imipenem | 0.2% (1) | 0.0% (0) | 0.18 | 2.5% (3) | 8.0% (2) | 0.43 |
| Meropenem | 0.0% (0) | 0.0% (0) | - | 3.3% (4) | 8.0% (2) | 0.86 |

*Klebsiella, Proteus spp, Pseudomona aeruginosa* and *Enterococcus faecalis* were more prevalent in men and Escherichia coli in women. Only enterobacteria behaved differently with respect to antibiotic resistance patterns, being higher in the male population.

In patients older than 65, a higher prevalence of *Enterococcus faecalis* was observed and there were no differences in antimicrobial susceptibility patterns. **(Table 6)**.

**Table 6.** Overall antibiotic resistance patterns for bacteria found in urine cultures of patients with community-acquired urinary tract infection admitted to HCUC or HSR, showing differences by sex and age group. It should be noted that bacteria classified as *other enterobacteria and **other bacteria were not considered for the calculation of % resistance. The following were considered: *Escherichia coli, Klebsiella spp, Enterococcus faecalis, Enterococcus faecium, Proteus spp, Pseudomonas aeruginosa, Enterobacter cloacae, Staphylococcus spp*.

| Antibiotic | Men;<br>% (n) | Women;<br>% (n) | p | ≥65 years<br>% (n) | <65 years<br>% (n) | p |
| --- | --- | --- | --- | --- | --- | --- |
| Cephalosporins 1<sup>st</sup> | 49.0% (121) | 28.4% (143) | <0.01 | 32.7% (144) | 38.7% (120) | 0.09 |
| Cephalosporins 2<sup>nd</sup> | 31.2% (77) | 14.1% (71) | <0.01 | 21.1% (93) | 17.7% (55) | 0.22 |
| Nitrofurantoin | 19.7% (59) | 10.8% (59) | <0.01 | 13.5% (68) | 14.6% (50) |  |
| Ciprofloxacin | 51.0% (154) | 29.1% (160) | <0.01 | 35.9% (184) | 38.2% (130) |  |
| Gentamicin | 21.2% (59) | 12.2% (63) | <0.01 | 15.4% (73) | 15.2% (49) |  |
| Trimethoprim-Sulfamethoxazole | 38.3% (101) | 31.9% (163) | 0.05 | 33.6% (153) | 34.7% (111) |  |
| Cephalosporins 3<sup>rd</sup> | 42.7% (114) | 19.1% (98) | <0.01 | 28.1% (130) | 25.9% (82) | 0.32 |
| Piperacillin-Tazobactam | 8.6% (23) | 2.5% (13) | <0.01 | 4.5% (21) | 4.7% (15) | 0.92 |
| Ertapenem | 9.1% (23) | 1.8% (9) | <0.01 | 3.6% (16) | 5.1% (16) | 0.31 |
| Imipenem | 3.4% (9) | 1.0% (5) | 0.01 | 2.2% (10) | 1.3% (4) | 0.32 |
| Meropenem | 3.0% (8) | 0.2% (1) | <0.01 | 1.3% (6) | 0.9% (3) | 0.61 |
| Amikacin | 4.9% (13) | 0.2% (1) | <0.01 | 1.5% (7) | 2.2% (7) | 0.51 |
| Oxacillin | 36.4% (4) | 0.0% (0) | 0.45 | 30.0% (3) | 16.7% (1) |  |
| Vancomycin | 4.3% (2) | 16.7% (7) | 0.12 | 8.5% (5) | 13.8% (4) |  |
| Teicoplanin | 0.0% (0) | 2.7% (1) | - | 2.0% (1) | 0.0% (0) |  |
| Linezolid | 0.0% (0) | 0.0% (0) | - | 0.0% (0) | 0.0% (0) |  |

## DISCUSSION

CA-UTI is a highly prevalent pathology and a frequent reason for hospital admission in the adult population (19–21). One of the factors that can determine morbidity and mortality is the correct initial empirical antibiotic choice, especially in patients who require hospitalization. To assist in the latter, it is necessary to identify antimicrobial susceptibility patterns. However, information regarding patients who require hospitalization has been poorly represented in available reports.

Our study recruited more than 800 patients requiring hospitalization with CA-UTI at two hospitals/health centers. The majority were women with ages close to 65. About 70% of the patients had more than one associated condition, as evidenced in previous reports, and presented bacteria resistant to first-line antibiotics. The most frequent causative agents were enterobacteria, *Escherichia coli* and *Klebsiella spp*. (68.1% and 17.7%, respectively). Similar to other reports, the presence of Pseudomonas, Proteus, Enterobacter cloacae and gram-positive bacteria was infrequent (8,22).

While it is true that the types and frequencies of bacteria detected are similar to those reported in the literature, there are differences with respect to antimicrobial susceptibility patterns. Overall, our study detected a high prevalence of bacteria resistant to first-line antibiotics. Approximately 30% of the enterobacteria were resistant to 1st generation cephalosporins, quinolones and trimethoprim-sulfamethoxazole, whereas 20% were resistant to 2nd and 3rd generation cephalosporins. We observed an opposite situation for piperacillin-tazobactam, carbapenems and amikacin, which presented overall resistance rates below 5%, making them good empirical therapeutic options. The same concept can be applicable to vancomycin when infection by gram-positive bacteria is suspected, given resistance rates of less than 11% (Table 4).

When analyzing the susceptibility patterns of each of the bacteria, we observed that the resistance rates of *Escherichia coli* strains to 1st generation cephalosporins, ciprofloxacin and trimethoprim-sulfamethoxazole exceeded 30%. For 2nd and 3rd generation cephalosporins, resistance rates were lower, but above the thresholds suggested by clinical guidelines for their empirical use as 1st line antibiotics. On the other hand, amikacin, carbapenems and piperacillin-tazobactam showed resistance rates below 1%. Previous studies have reported mixed results. In Chile, the latest report from the Bacterial Resistance Collaborative Group (2012) reported lower resistance of *Escherichia coli* to ciprofloxacin and ceftriaxone (22.8% and 7.8%, respectively) in adult ambulatory urine cultures, but similar rates among 1st generation cephalosporins, nitrofurantoin and amikacin (37.6%, 4.7% and 0.1%). Although these data are not completely adaptable to the type of patients included in this study, our results show a clear increase in the rate of bacterial resistance for 3rd generation cephalosporins (23).

Studies that included patients similar to ours (hospitalized) have shown disparate results. Jia et al. (24) reported higher resistance rates in Asian patients for ciprofloxacin (50.2%), trimethoprim-sulfamethoxazole (55.6%), as well as 2nd and 3rd generation cephalosporins (38%). However, there was a similarity regarding resistance for amikacin, nitrofurantoin and carbapenems. Mitcho et al. in Poland reported much lower resistance rates for E. coli for 3rd generation cephalosporins (13%), but higher for amikacin (11.8%) and piperacillin-tazobactam (23%) (22). Gajdács et al. also reported low resistance rates for 3rd generation cephalosporins (13.1%) in older Hungarian adults, and were similar to our findings for the rest of the antibiotics. It should be noted that, in this study, the authors did not define whether the UTIs were acquired before or during the hospital stay (25).

Regarding the second most prevalent bacterium (i.e., *Klebsiella spp*), its strains presented the highest resistance rates for all antibiotics, with only amikacin, imipenem and meropenem suggested as good empirical therapeutic options. Similar results were reported by Hrbacek et al. in hospitalized patients, differing only for piperacillin-tazobactam, where resistance was much lower (17%) (26). Gajdács et al. detected differences only for trimethoprim-sulfamethoxazole and 3rd generation cephalosporins, which had lower resistance rates than ours (27.8% and 27.7%, respectively) (25). It should be noted that in both studies it was not possible to discern whether the cultures corresponded to community-acquired or nosocomial infections.

Although infections by gram-positive bacteria were observed at low frequencies, *Enterococcus spp*. ranked third with a rate of 8.6% (6.7% for *E. faecalis* and 1.9% for *E. faecium*). Resistance to ampicillin was null for *E. faecalis*, but very high for *E. faecium*. Something similar occurred for nitrofurantoin (0% and 68%) and vancomycin (0% and 56.3%). It is noteworthy that resistance to quinolones was high for both *Enterococcus*, being higher for *E. faecium*. The resistance patterns detected by Hrbacek et al. and Gajdács et al. were similar. However, their data combined both types of *Enterococcus* (25,26).

Our study has some limitations. Few patients belonging to the public health care center were included, which could affect the extrapolation of the results to the entire population. On the other hand, the susceptibility of some currently available antibiotics, such as Fosfomycin and new cephalosporins associated with beta-lactamase inhibitors, was not studied.

Among the main strengths of our study is the presence of a spectrum of patients of high frequency and clinical relevance, who have been underrepresented in published studies, including patients requiring hospitalization for a community-acquired infection. Also noteworthy is the prospective nature and the recruitment of a large number of patients.

Our data show that resistance rates for the most frequently-used antibiotics in our country are well above the recommended threshold for the selection of empirically-founded antibiotics. It is suggested here that the use of quinolones (ciprofloxacin) and cephalosporins could be replaced by amikacin. This switch represents further benefits in relation to its low cost and easy administration. In view of the latter, we believe that the recommendations of the current guidelines should be reviewed and updated.

Finally, our results emphasize the importance of having local information on antimicrobial susceptibility, especially for patients who develop community infections with risk factors associated to hospital-acquired infections. These results can offer medical teams more knowledge and tools to select appropriate antibiotic treatments before obtaining urine cultures and antibiograms, thus improving morbimortality and length of stay of patients with CA-UTI.

## Data Availability

All relevant data are within the manuscript and its Supporting Information files.

## ACKNOWLEDGMENTS

We thank Carolina Henriquez for her excellent logistic support during the execution of the study. Thanks to Camila Marti, Jose Maria Garcia, Miguel Cantillana, Jennifer Vilo and Mayerling Malig for their support in patient recruitment, as well as to Dr. Patricia García for her technical assistance from the clinical laboratory service

## Notes

### Competing Interest Statement

The authors have declared no competing interest.

### Funding Statement

The authors received no specific funding for this work.

### Author Declarations

This study was approved by the Health Sciences Scientific Ethics Committee of the School of Medicine of the Pontifical Catholic University and the South-East Metropolitan Health Service. Number ID 2003005003. No consent was required given that the data were obtained and analyzed anonymously.

